# Antibody-independent microvascular inflammation impacts long-term risk in heart transplantation

**DOI:** 10.1101/2025.07.29.25332346

**Authors:** Shi Huang, Nelson Chow, Kyle Saysana, Eric Farber-Eger, Quinn S. Wells, David W. Bearl, JoAnn Lindenfeld, Kelly H. Schlendorf, Kaushik Amancherla

## Abstract

**Background:** Microvascular inflammation (MVI) following heart transplantation can occur with or without circulating anti-HLA donor-specific antibodies (DSAs). We sought to characterize the relationship between MVI, with or without accompanying DSA, and post-transplant outcomes.

**Methods:** We analyzed 8,305 endomyocardial biopsies (EMB) from 832 adult and pediatric HT recipients between July 1, 2013 and October 31, 2023. EMBs were graded by consensus guidelines, with MVI defined as pAMR grade ≥1. Rejection phenotypes were classified as no rejection, isolated cellular rejection (ACR), DSA-negative MVI, and DSA-positive MVI. Cox models with time-varying covariates were constructed to evaluate associations with incident CAV and mortality, adjusting for donor and recipient age.

**Results:** Among 832 HT recipients, 238 developed CAV and 121 died over a median follow-up of 4 years (IQR 2.3-6.4 years). Compared with individuals who never experienced biopsy-proven rejection, DSA-negative MVI was independently associated with CAV (HR, 1.47; 95% CI 1.01−2.16). DSA-positive MVI was associated with mortality (HR 1.97; 95% CI 1.07−3.64) with DSA-negative MVI demonstrating directional-concordance (HR 1.50, 95% CI 0.87-2.57), independent of CAV (HR 1.71, 95% CI 1.13-2.58). These associations remained consistent when stratified by adult and pediatric subgroups and in a six-month landmark sensitivity analysis.

**Conclusions:** MVI, with or without DSA, may be harmful in HT, extending recent renal findings to thoracic transplantation. Understanding the mechanistic basis for these results will be essential for identifying novel targets for therapeutic modulation and prolonging graft survival.

**HIGHLIGHTS:**

- Microvascular inflammation (representing pAMR ≥1) following heart transplantation is associated with increased risk of CAV and all-cause mortality, with or without accompanying anti-HLA donor-specific antibodies.
- Microvascular inflammation may serve as a modifiable target in post-transplant care.
- Urgent mechanistic studies are needed to distinguish between pathogenic vascular injury warranting intensified surveillance and immunosuppression, and clinically-inconsequential immune activity.

## INTRODUCTION

Repetitive alloimmune injury is a central driver of poor long-term survival after solid organ transplantation. Though “microvascular inflammation” (MVI)—inflammatory vascular injury with or without overt antibody-mediated rejection (AMR)—is associated with poor outcomes after renal transplantation^1^, its importance in thoracic transplantation where vascular injury represents a major life-limiting complication remains unclear. This uncertainty is clinically significant. Cardiac allograft vasculopathy (CAV)—the major barrier to long-term survival following heart transplantation (HT)^2^—originates from alloimmune vascular injury, which may be subclinical or histologically-subtle^2^. Current diagnostic criteria classify AMR-related injury by presence of MVI, emphasizing circulating donor-specific anti-HLA antibodies (DSAs)^3,4^, with therapies directed at reducing DSAs to limit graft injury. Yet, the extent to which MVI, particularly in DSA-negative settings, portends future CAV or graft failure remains unclear. Treatment decisions in this context remain individualized, with substantial institutional variation, owing to limited large-scale experience in tracking the relevance of MVI on outcomes^4^.

To address this gap, we leveraged a granular longitudinal transplant database at a high-volume transplant center to evaluate whether biopsy-defined MVI, with or without accompanying DSA, is associated with adverse post-transplant outcomes. Specifically, and analogous to renal transplantation, we tested the hypothesis that DSA-negative MVI is independently associated with development of CAV, providing a histologic signal of alloimmune injury that may extend beyond conventional serologic definitions.

## METHODS

### Study Cohort

We conducted a retrospective cohort study of all adults and children who underwent heart transplantation (HT) at Vanderbilt University Medical Center (VUMC) between July 1, 2013 and October 31, 2023. This time frame was intentionally selected for two reasons: **(1)** the International Society for Heart and Lung Transplantation (ISHLT) Working Formulation^5^ for diagnosis of antibody-mediated rejection (AMR) was published in 2013 and our center began routinely reporting both histological evidence of AMR (pAMR1-h) and immunological evidence (pAMR1-I, based on C4d deposition) on biopsies; and **(2)** the time window allowed for adequate longitudinal follow-up for coronary surveillance to assess cardiac allograft vasculopathy (CAV) status.

Adult HT recipients received induction therapy with corticosteroids alone or in combination with basiliximab or anti-thymocyte antibody (thymoglobulin), followed by maintenance immunosuppression consisting of calcineurin inhibitor, anti-proliferative agent (e.g., mycophenolate mofetil), and prednisone taper per institutional protocol with planned discontinuation by 4-6 months post-HT. Pediatric HT recipients uniformly received thymoglobulin for induction, followed by a 5-day steroid taper. Maintenance immunosuppression in children included a calcineurin inhibitor and antiproliferative agent for the first 3 months post-HT, with an mTOR inhibitor replacing the antiproliferative agent subsequently.

Surveillance for circulating donor-specific anti-HLA antibodies (DSAs) was conducted at post-HT weeks 2, 4, 12, and 24, followed by at least once every 6 months or as clinically indicated. Per institutional protocol in adults, surveillance endomyocardial biopsies (EMBs) were performed coinciding with the steroid taper and were typically performed on post-HT weeks 2, 4, 8, 12, 16, and 20 or as clinically indicated. Baseline coronary angiography typically occurred between 2-3 months post-HT with annual angiographic surveillance initiated at 1-year post-HT. This study was approved by the VUMC Institutional Review Board (#200551).

#### Exclusion criteria

1. HT recipients transplanted at other centers and who transferred care to VUMC without verifiable EMB data;
2. Veterans Affairs (VA) patients who underwent HT at VUMC but received post-transplant care exclusively within the VA system;
3. Absence of coronary angiography data for CAV surveillance.

### Data Collection

Recipient data was extracted from VUMC’s electronic health record and donor data was abstracted from the United Network for Organ Sharing (UNOS) database. Recipient variables included age at transplant, sex, comorbidities, and DSAs. Donor variables included donor age and method of organ procurement (donation after brain death [DBD] or circulatory death [DCD]).

### Rejection Grading

Following EMB collection and processing, sections were transferred to the pathology department in formalin or in saline. Formalin-fixed, paraffin-embedded specimens underwent sectioning, followed by hematoxylin and eosin (H&E) staining. Specimens transferred in saline were embedded in optimal cutting temperature (OCT) compound and underwent sectioning and immunofluorescence staining for C4d.

All EMB specimens were graded by cardiac pathologists with expertise in heart transplantation. Acute cellular rejection (ACR) was graded according to ISHLT criteria^6^, with ACR grade 2R and 3R considered clinically significant rejection. Antibody-mediated rejection (AMR) was graded according to ISHLT consensus^5^. Histologic evidence of AMR (pAMR1-h) was characterized by endothelial swelling in the absence of peri-capillary C4d deposition.

Immunological evidence of AMR (pAMR1-i) was characterized by peri-capillary C4d deposition in >50% of capillaries (excluding peri-myocyte staining). The presence of both histologic and immunologic evidence of AMR was defined as pAMR2, with pAMR3 representing severe AMR with concomitant hemodynamic compromise^3^.

For primary analytic purposes, MVI was defined as the presence of pAMR1-h, pAMR1-i, or pAMR2/3. MVI was further stratified as DSA-positive or DSA-negative based on serologic status at the time of biopsy. Individuals who never demonstrated biopsy-proven rejection over the follow-up period comprised the “No rejection” group while those with ACR only (without MVI or AMR) were classified as “ACR only”. Of note, per institutional protocol, adults with pAMR1-i, pAMR2, or pAMR3 are treated as definitive AMR with therapies targeted at reducing DSA. On the other hand, pAMR1-h typically does not result in intensified immunosuppression (unless other markers of graft injury, such as graft dysfunction, are present) but, rather, is closely followed for resolution.

CAV surveillance was performed by invasive coronary angiography, considered the gold standard by current ISHLT consensus^2^. In a small number of cases where individuals underwent redo HT or autopsy, CAV grading was adjudicated by direct pathological examination of the coronaries. Briefly, CAV-1 represents mild angiographically-evident disease in the coronary vasculature, CAV-2 denotes ≥70% in a single major vessel or severe branch disease in two coronary distributions, and CAV-3 includes left main coronary disease or severe multivessel involvement^2^. Additionally, CAV-3 could be diagnosed in the context of graft dysfunction with underlying CAV-1 or CAV-2^2^. Clinically significant CAV was considered CAV grade ≥1, which is associated with increased risk of graft loss and mortality after transplantation^2,7–9^.

### Outcomes

The primary outcomes were incident CAV and all-cause mortality.

### Statistical analysis

Continuous variables were summarized as median (interquartile range [IQR]) and categorical variables were described as N (%). To account for time-varying exposures, Cox proportional hazards models were constructed with rejection phenotype and DSA status treated as time-varying covariates. In the mortality analysis, CAV was also treated as a time-varying covariate. DSA status was also modeled as a separate variable, as it may be present independently of rejection and can fluctuate over time irrespective of biopsy findings. This approach mitigates immortal time bias and allows for granular alignment of evolving immunologic status (e.g., rejection states, presence or absence of circulating DSA, etc.) with individual-level risk. Rejection status and DSA exposure were updated at each biopsy, and person-time was recalculated accordingly. The CAV outcome was based on last date of coronary evaluation^2^. Patients without CAV were censored at their last coronary interrogation.

Adjusted hazard ratios (HR) and their 95% confidence intervals (CI) are reported. Kaplan–Meier curves were constructed to visualize unadjusted time-to-event distributions. A six-month landmark sensitivity analysis was conducted to address early post-HT immune dynamics and surveillance intensity and to assess robustness of our primary model. Within the landmark analysis, rejection phenotypes were categorized into their most severe rejection episode during the six-month post-HT period (hierarchically: DSA-positive MVI > DSA-negative MVI > ACR only > no rejection). All analyses were performed in R version 4.4.0 (R Core Team).

## RESULTS

We identified 832 adults and children who underwent heart transplantation between July 1, 2013 and October 31, 2023. The median recipient age at HT was 51 years (IQR 33-62 years) and 273 (33%) of recipients were female. The median donor age was 28 years (IQR 19-36 years). In total, 8,305 EMBs were included in the analysis, spanning the spectrum of rejection phenotypes (**Figure 1**). The first EMB post-HT was performed at a median of 11 days (IQR 7-15 days). Among the cohort, 287 individuals (34%) developed circulating DSAs, first detected at a median of 187 days (IQR 16-720 days) post-HT. Full demographics are outlined in **Table 1**. **Figure 2A** and **2B** depict the Kaplan-Meier curves for incident CAV and all-cause mortality for the entire cohort.

**Figure 1.**
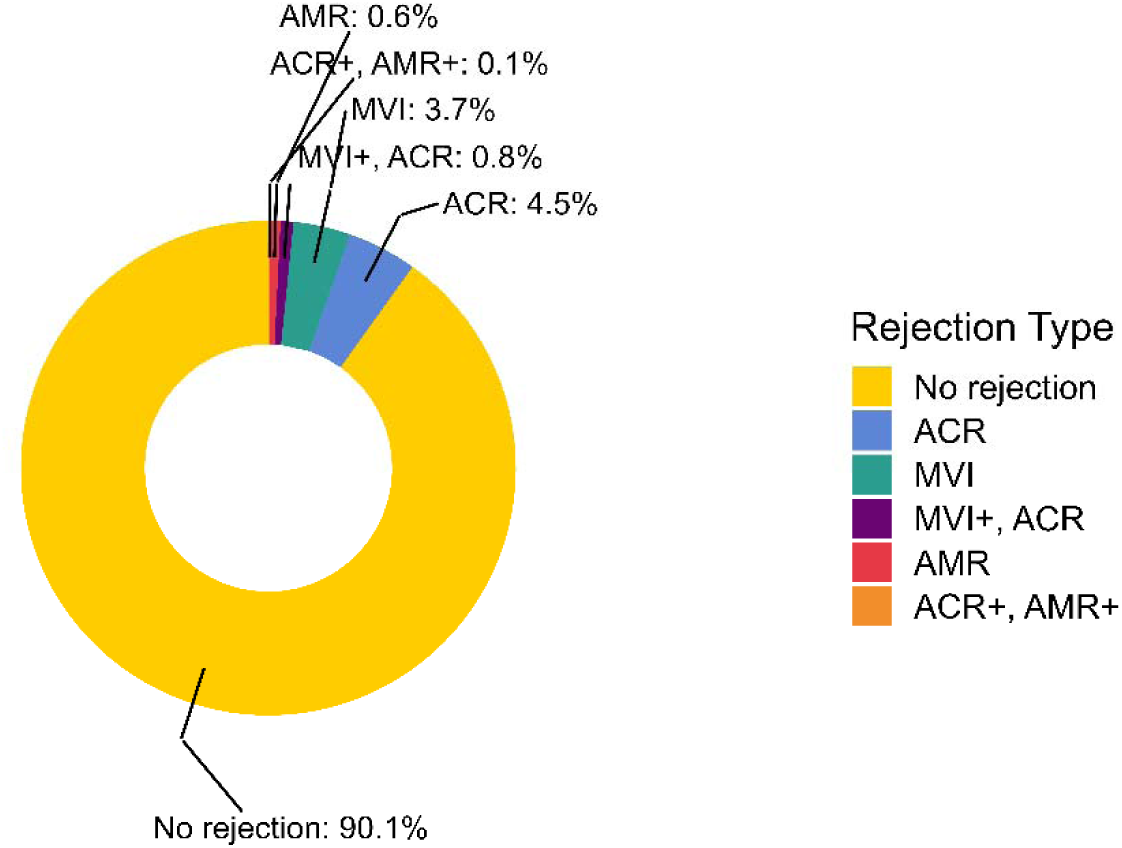
Distribution of rejection phenotypes across 8,305 endomyocardial biopsies. ACR = acute cellular rejection; MVI = microvascular inflammation (defined here as pAMR1-h or pAMR1-i); AMR = antibody-mediated rejection (defined here are pAMR2 or pAMR3).

**Figure 2.**
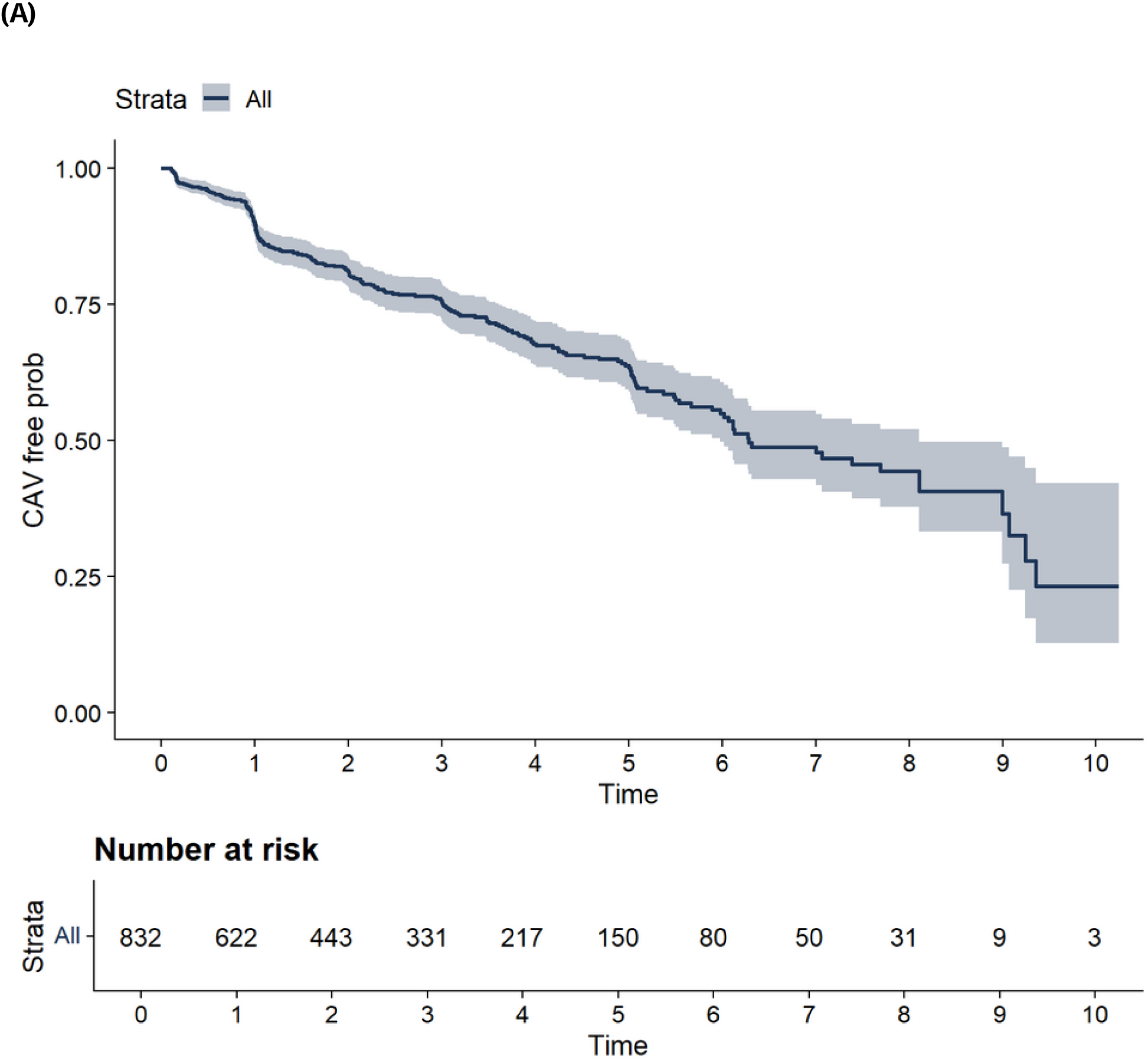
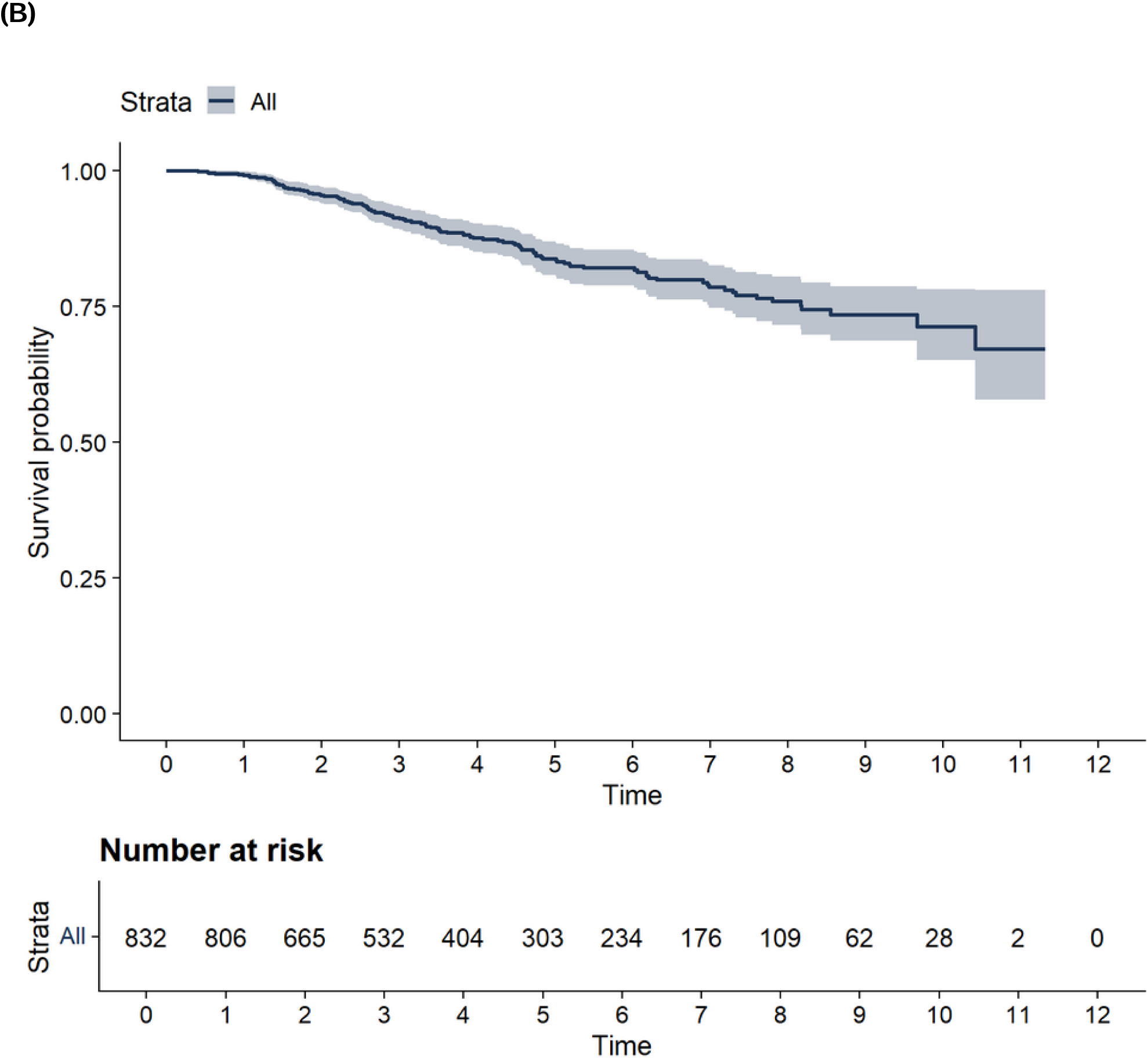
CAV-free survival and all-cause mortality following heart transplantation. (**A**) Kaplan-Meier curve for CAV-free survival following transplantation in the entire cohort. Individuals were censored at last coronary assessment (e.g., angiography, pathology of explanted heart at time of re-do transplant or autopsy). **(B)** Kaplan-Meier curve for all-cause mortality following transplantation in the entire cohort.

**Table 1.** Baseline cohort demographics.

| Characteristic | N = 832<sup>1</sup> |
| --- | --- |
| Recipient age at transplant (years) | 51 (33, 62) |
| Recipient sex (Female) | 273 (33%) |
| Donation type |  |
| Circulatory death | 135 (16%) |
| Brain death | 697 (84%) |
| Donor age (years)* | 28 (19, 36) |
| Days to first biopsy | 11 (7, 15) |
| Died during follow-up | 121 (15%) |
| Cardiac allograft vasculopathy | 238 (29%) |
| Donor-specific antibodies | 287 (34%) |
| <sup>1</sup> Median (Q1, Q3); n (%) |  |
| *N = 831 |  |

We first examined outcomes among individuals who experienced pAMR1 (defined as either pAMR1-h or pAMR1-i^3,5^) during follow-up (**Figure 3A**). Among 154 individuals who experienced pAMR1 without progression to pAMR2, 63 (40.9%) developed CAV. An additional 10 individuals progressed from pAMR1 to pAMR2, with 3 (30.0%) subsequently developing CAV. In contrast, among those who experienced pAMR2 without prior pAMR1 (n = 9), 5 individuals (55.6%) developed CAV. Notably, 4 of 15 individuals (26.7%) who experienced pAMR2 followed by a later episode of pAMR1 also developed CAV. These findings suggest that MVI—captured at the pAMR1 stage—may represent an early and clinically meaningful precursor to vascular injury, even in the absence of progression to pAMR2/3. For comparison, 40 of 123 individuals (32.5%) in the “ACR only” group developed CAV and 123 of 521 (23.6%) recipients with no biopsy-proven rejection during follow-up developed CAV. These findings support the clinical relevance of MVI, particularly in the form of isolated pAMR1, as a high-risk phenotype associated with increased CAV risk. While the number of individuals with sequential rejection phenotypes was small, the high event rate among those with isolated pAMR2 further underscores the pathogenic potential of sustained or severe vascular injury. Together, these results suggest that MVI—whether transient or persistent—may be an early and meaningful contributor to CAV, even in the absence of concurrent DSA or progression to more advanced AMR.

**Figure 3.**
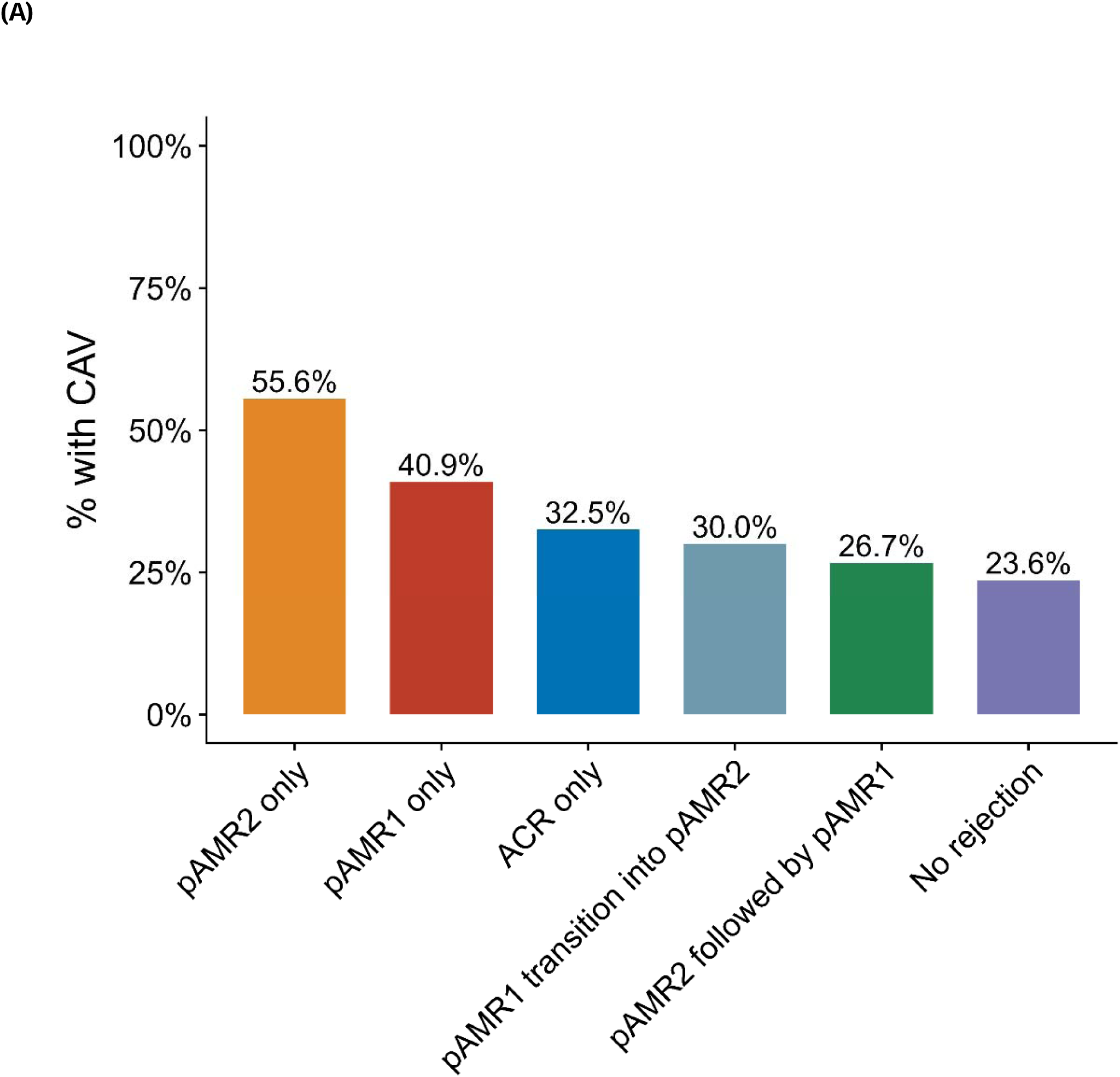
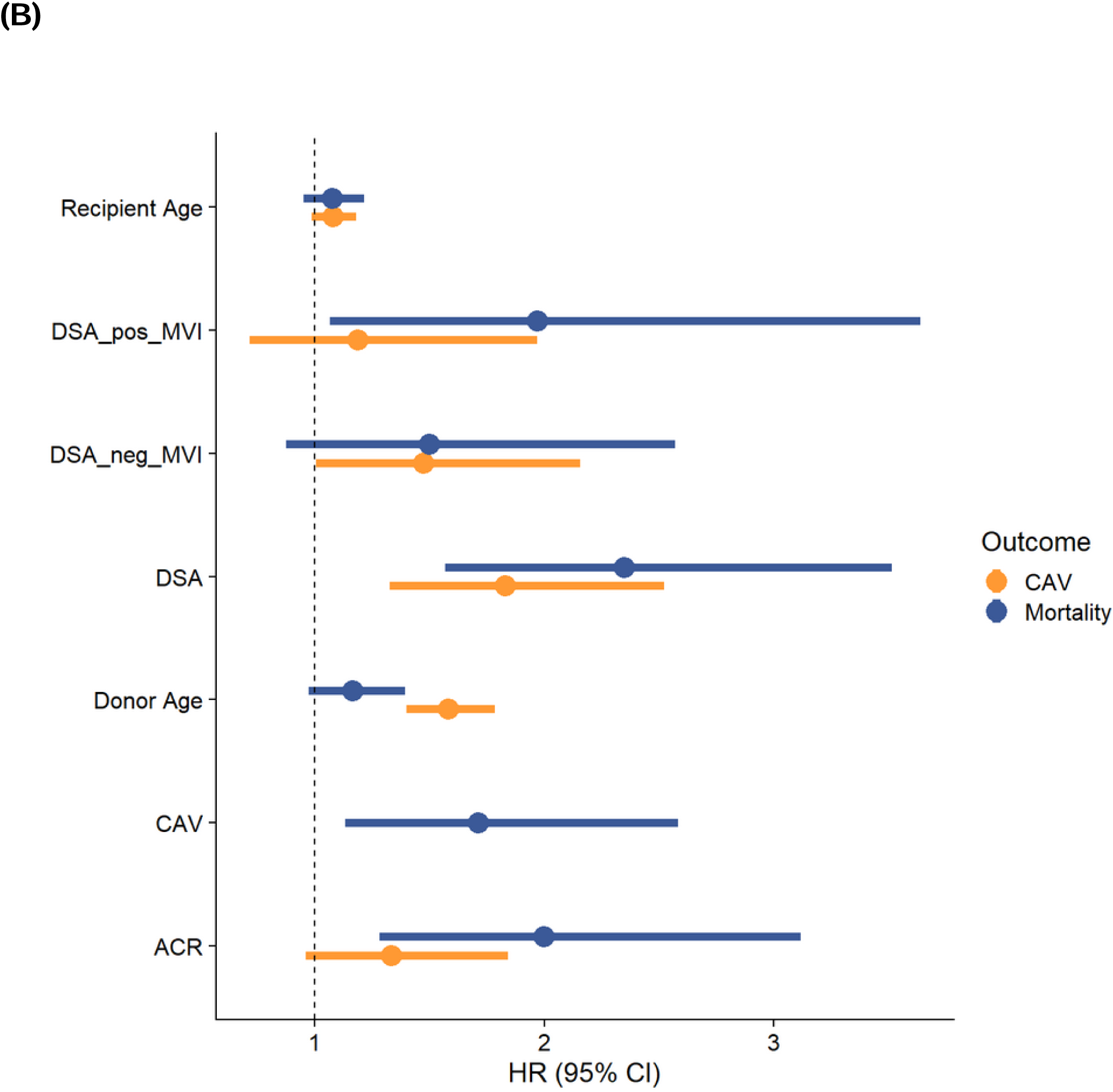
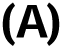
Association between microvascular inflammation (MVI), cardiac allograft vasculopathy (CAV), and all-cause mortality. **(A)** Percentage of heart transplant (HT) recipients with varying rejection phenotypes—diagnosed on endomyocardial biopsy—who subsequently developed CAV. **(B)** Forest plot of hazard ratios (HRs) for incident CAV (yellow) and all-cause mortality (blue) derived from Cox proportional hazard models. In the CAV analysis, rejection phenotype and circulating DSAs were treated as time-varying covariates, while donor and recipient age were treated as fixed covariates. In the mortality analysis, CAV— the leading case of late mortality after HT—was also treated as a time-varying covariate.

Over a median 4-year follow-up (IQR 2.3-6.4 years), 238 recipients developed CAV and 121 died. Among 154 individuals with pAMR1, 63 (40.9%) developed CAV compared to 23.6% of those without rejection (**Figure 1A**). In comparison to “no rejection”, DSA-negative MVI was independently associated with CAV (hazard ratio [HR], 1.47; 95% confidence interval [CI] 1.01– 2.16; **Figure 1B**). DSA-positive MVI was associated with mortality (HR 1.97; 95% CI 1.07–3.64) with DSA-negative MVI demonstrating directional-concordance (HR 1.50, 95% CI 0.87-2.57), independent of CAV (HR 1.71, 95% CI 1.13-2.58).

We also conducted several pre-specified subgroup and sensitivity analyses. First, we stratified the cohort by age to evaluate whether the associations observed in the overall cohort were consistent across adult (N = 693) and pediatric (N = 139) HT recipients. In adults, both the DSA-negative MVI (HR 1.39, 95% CI 0.94-2.06) and DSA-positive MVI (HR 1.11, 95% CI 0.65-1.89) phenotypes demonstrated directional concordance with the primary analyses regarding their association with incident CAV (**Supplemental Figure 1A**). Similar trends were observed for mortality. In pediatric HT recipients, the relationship between MVI and CAV was also directionally consistent with the overall cohort (**Supplemental Figure 1B**). However, precision was limited in this subgroup due to the smaller sample size and fewer CAV events, resulting in wider confidence intervals.

We further performed a six-month landmark sensitivity analysis to account for early post-HT surveillance intensity and possible immortal time bias (**Supplemental Figure 2**). This analysis demonstrated results that were directionally aligned with our primary time-varying models. These findings suggest that early histopathologic evidence of microvascular injury may set the trajectory for later CAV development.

## DISCUSSION

In this large, granular, longitudinal cohort of HT recipients, we found that MVI—whether or not accompanied by circulating anti-HLA DSAs—was associated with adverse long-term outcomes, including CAV and all-cause mortality. These findings extend observations from renal transplantation to thoracic transplantation and suggest that subclinical alloimmune microvascular injury may represent a shared mechanism of chronic graft dysfunction across organ systems^1^. Importantly, DSA-negative MVI was not benign, but rather, it conferred a significant risk of CAV development, independent of histologic ACR or circulating DSA.

These results are particularly informative in light of the recent consensus report on AMR^4^, which explicitly outlines the need for improved understanding of DSA and MVI. Further, the consensus statement emphasizes that center-level practice variation remains substantial, particularly in the surveillance, reporting, and treatment of MVI and AMR, often driven by individual protocols and local interpretations. Despite the widespread use of ISHLT grading systems^4–6^, the document acknowledges that therapeutic decision-making in pAMR1 remains highly inconsistent, and that robust, outcome-based data are lacking to guide standardized care in this group.

Our real-world findings help fill this critical knowledge gap, with our results indicating that MVI—regardless of DSA status—identifies patients at increased risk of CAV. In our cohort, 154 individuals experienced pAMR1 (either histologic [pAMR1-h] or immunologic [pAMR1-i]) without progression to pAMR2 and 63 of these (40.9%) subsequently developed CAV. An additional 10 individuals progressed from pAMR1 to pAMR2, with 3 (30.0%) developing CAV. Among those with isolated pAMR2 (n = 9), 5 (55.6%) developed CAV, while 4 of 15 (26.7%) who experienced pAMR2 followed by later pAMR1 also developed CAV. These findings strongly reinforce the hypothesis that MVI—captured at the pAMR1 stage—may serve as a critical early immunopathologic trigger for vascular injury, even when formal criteria for AMR are not met. By contrast, the incidence of CAV was 32.5% among those with ACR only and 23.6% among individuals who never demonstrated histologic rejection, highlighting the relative risk gradient conferred by MVI.

The progressive nature of MVI and its potential to evolve into more severe AMR raises important clinical considerations. First, these findings support the concept that pAMR1 may not represent a static phenotype but rather a transitional state within the continuum of alloimmune injury. Second, the observation that ≈41% of individuals with pAMR1 (in the absence of pAMR2) developed CAV underscores the need to re-evaluate whether isolated MVI—regardless of DSA or graft dysfunction—warrants intensified surveillance or preemptive therapeutic intervention.

This is especially relevant in light of the most recent ISHLT consensus^4^, which recommends treatment primarily in cases of pAMR2/3 while offering limited guidance on the management of pAMR1. Our findings directly challenge the prevailing assumption that pAMR1, particularly when DSA-negative, is benign and instead suggest that earlier recognition and management of MVI may be critical to improving long-term outcomes after heart transplantation. As clinical practice continues to evolve, these data provide real-world support for incorporating MVI more formally into post-HT risk stratification algorithms.

These observations also reinforce the potential for MVI to serve as a modifiable target in post-HT care. As mechanistic studies emerge—particularly those leveraging single-cell and spatial transcriptomic profiling to delineate cell-type–specific signatures of microvascular injury^10,11^— there is growing opportunity to discern transient, clinically-inconsequential immune activity from sustained pathogenic microvascular injury. In parallel, the role of non-HLA antibodies (e.g., anti-angiotensin II type 1 receptor antibodies)^12^, natural killer (NK) cell– mediated cytotoxicity^13^, and complement-independent endothelial injury are increasingly recognized as potential drivers of MVI, independent of conventional AMR. Dissecting these overlapping yet distinct immunologic axes will be essential for developing rational, targeted interventions aimed at mitigating progressive vascular injury and improving long-term graft outcomes. Our study provides a foundation upon which these mechanistic layers can be superimposed.

### Study limitations

This study represents a large, real-world experience in HT, incorporating granular, longitudinal rejection phenotyping and angiographic CAV surveillance. While our approach enabled a detailed characterization of temporal alloimmune microvascular injury and its relationship with long-term vascular outcomes in both adult and pediatric HT recipients, several limitations merit consideration. As with any observational cohort, findings are subject to residual confounding and unmeasured bias despite our application of time-varying methods that account for dynamic changes in rejection phenotypes and exposure status. The diagnosis of MVI and its classification are based on histopathologic criteria and may be affected by interobserver variability^14^. As rejection phenotypes were adjudicated by a dedicated group of cardiac pathologists at our institution, interobserver variability in the diagnosis and grading of rejection phenotypes is attenuated. We further mitigated misclassification bias by using rigorous definitions—for both rejection and CAV grading—aligned with ISHLT consensus^2,3,5,6^. While our pediatric subgroup represents one of the largest modern cohorts evaluated in this context, the relatively low event rate in this population limits statistical power and the precision of risk estimates. However, the directionality of observed associations was consistent across age strata, supporting the biological plausibility of our findings. Finally, as a single-center study, the generalizability of our findings may be influenced by institution-specific immunosuppression protocols, biopsy schedules, and DSA detection thresholds. Nevertheless, our consistent surveillance strategies, including angiographic CAV assessment by experienced interventional and transplant cardiologists and histopathologic evaluation by a dedicated team of cardiac pathologists, strengthen internal validity and support the reproducibility of our analytic framework for future multicenter validation efforts.

### Conclusions

Our observations suggest MVI may be harmful in HT (with or without DSA), extending recent renal findings to thoracic transplantation^1^. Although observational confounding (e.g., selection bias, biopsy frequency) are important limitations, our results reflect a large-scale real-world experience that underscores the prognostic import of MVI currently not widely treated^4^.

This may be especially crucial in thoracic transplants, where progressive graft failure presaged by MVI is often catastrophic and irreversible. With novel technologies that allow access to patient-based heterogeneity in rejection at cellular resolution, understanding the mechanistic basis for this observation—and potential therapeutic routes (including beyond conventional HLA-directed therapies)—will be critical to extending graft survival across solid organ transplantation.

## Data Availability

All data produced in the present study are available upon reasonable request to the authors.

## REFERENCES

1. Sablik M, Sannier A, Raynaud M, Goutaudier V, Divard G, Astor BC, Weng P, Smith J, Garro R, Warady BA, et al. Microvascular Inflammation of Kidney Allografts and Clinical Outcomes. N Engl J Med. 2025;392:763–776. doi: 10.1056/NEJMoa2408835

2. Mehra MR, Crespo-Leiro MG, Dipchand A, Ensminger SM, Hiemann NE, Kobashigawa JA, Madsen J, Parameshwar J, Starling RC, Uber PA. International Society for Heart and Lung Transplantation working formulation of a standardized nomenclature for cardiac allograft vasculopathy-2010. J Heart Lung Transplant. 2010;29:717–727. doi: 10.1016/j.healun.2010.05.017

3. Colvin MM, Cook JL, Chang P, Francis G, Hsu DT, Kiernan MS, Kobashigawa JA, Lindenfeld J, Masri SC, Miller D, et al. Antibody-mediated rejection in cardiac transplantation: emerging knowledge in diagnosis and management: a scientific statement from the American Heart Association. Circulation. 2015;131:1608–1639. doi: 10.1161/CIR.0000000000000093

4. Kobashigawa J, Zuckermann A, Zeevi A, Barten MJ, Chang PP, Colvin M, Coutance G, Dipchand A, Ensminger S, Farrero M, et al. Summary of the International Society for Heart and Lung Transplantation consensus conference on emerging understanding of antibodies and antibody-mediated rejection in heart transplantation. J Heart Lung Transplant. 2025. doi: 10.1016/j.healun.2025.02.1690

5. Berry GJ, Burke MM, Andersen C, Bruneval P, Fedrigo M, Fishbein MC, Goddard M, Hammond EH, Leone O, Marboe C, et al. The 2013 International Society for Heart and Lung Transplantation Working Formulation for the standardization of nomenclature in the pathologic diagnosis of antibody-mediated rejection in heart transplantation. J Heart Lung Transplant. 2013;32:1147–1162. doi: 10.1016/j.healun.2013.08.011

6. Stewart S, Winters GL, Fishbein MC, Tazelaar HD, Kobashigawa J, Abrams J, Andersen CB, Angelini A, Berry GJ, Burke MM, et al. Revision of the 1990 working formulation for the standardization of nomenclature in the diagnosis of heart rejection. J Heart Lung Transplant. 2005;24:1710–1720. doi: 10.1016/j.healun.2005.03.019

7. Tremblay-Gravel M, Racine N, de Denus S, Ducharme A, Pelletier GB, Giraldeau G, Liszkowski M, Parent MC, Carrier M, Fortier A, White M. Changes in Outcomes of Cardiac Allograft Vasculopathy Over 30 Years Following Heart Transplantation. JACC Heart Fail. 2017;5:891–901. doi: 10.1016/j.jchf.2017.09.014

8. Prasad N, Harris E, Yuzefpolskaya M, DeFilippis EM, Colombo PC, Sayer G, Chernovolenko M, Fried J, Bae D, Oh KT, et al. Can the grading of mild cardiac allograft vasculopathy be further refined? An angiographic and physiologic assessment of heart transplant recipients with ISHLT CAV 1. J Heart Lung Transplant. 2025;44:905–912. doi: 10.1016/j.healun.2024.12.013

9. Chih S, Chong AY, Mielniczuk LM, Bhatt DL, Beanlands RS. Allograft Vasculopathy: The Achilles’ Heel of Heart Transplantation. J Am Coll Cardiol. 2016;68:80–91. doi: 10.1016/j.jacc.2016.04.033

10. Amancherla K, Qin J, Hulke ML, Pfeiffer RD, Agrawal V, Sheng Q, Xu Y, Schlendorf KH, Lindenfeld J, Shah RV, et al. Single-Nuclear RNA Sequencing of Endomyocardial Biopsies Identifies Persistence of Donor-Recipient Chimerism With Distinct Signatures in Severe Cardiac Allograft Vasculopathy. Circ Heart Fail. 2023;16:e010119. doi: 10.1161/CIRCHEARTFAILURE.122.010119

11. Amancherla K, Taravella Oill AM, Bledsoe X, Williams AL, Chow N, Zhao S, Sheng Q, Bearl DW, Hoffman RD, Menachem JN, et al. Dynamic responses to rejection in the transplanted human heart revealed through spatial transcriptomics. bioRxiv. 2025. doi: 10.1101/2025.02.28.640852

12. Butler CL, Hickey MJ, Jiang N, Zheng Y, Gjertson D, Zhang Q, Rao P, Fishbein GA, Cadeiras M, Deng MC, et al. Discovery of non-HLA antibodies associated with cardiac allograft rejection and development and validation of a non-HLA antigen multiplex panel: From bench to bedside. Am J Transplant. 2020;20:2768–2780. doi: 10.1111/ajt.15863

13. Loupy A, Duong Van Huyen JP, Hidalgo L, Reeve J, Racape M, Aubert O, Venner JM, Falmuski K, Bories MC, Beuscart T, et al. Gene Expression Profiling for the Identification and Classification of Antibody-Mediated Heart Rejection. Circulation. 2017;135:917–935. doi: 10.1161/CIRCULATIONAHA.116.022907

14. Crespo-Leiro MG, Zuckermann A, Bara C, Mohacsi P, Schulz U, Boyle A, Ross HJ, Parameshwar J, Zakliczynski M, Fiocchi R, et al. Concordance among pathologists in the second Cardiac Allograft Rejection Gene Expression Observational Study (CARGO II). Transplantation. 2012;94:1172–1177. doi: 10.1097/TP.0b013e31826e19e2

