## Supplemental Material for "Antibody-independent microvascular inflammation impacts long-term risk in heart transplantation"

**Supplemental Figure 1.** Time-varying Cox regression results of rejection phenotypes, CAV, and mortality in adults **(A**; N = 693**)** and children **(B**; N = 139**)** following heart transplantation.

**A.**

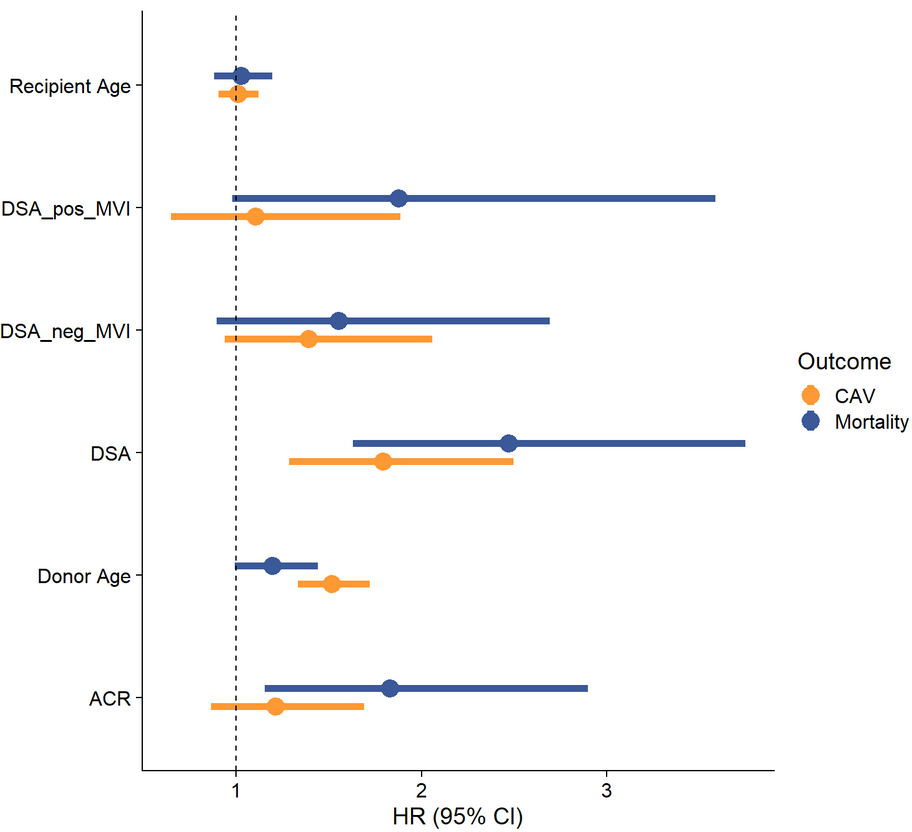

**B.**

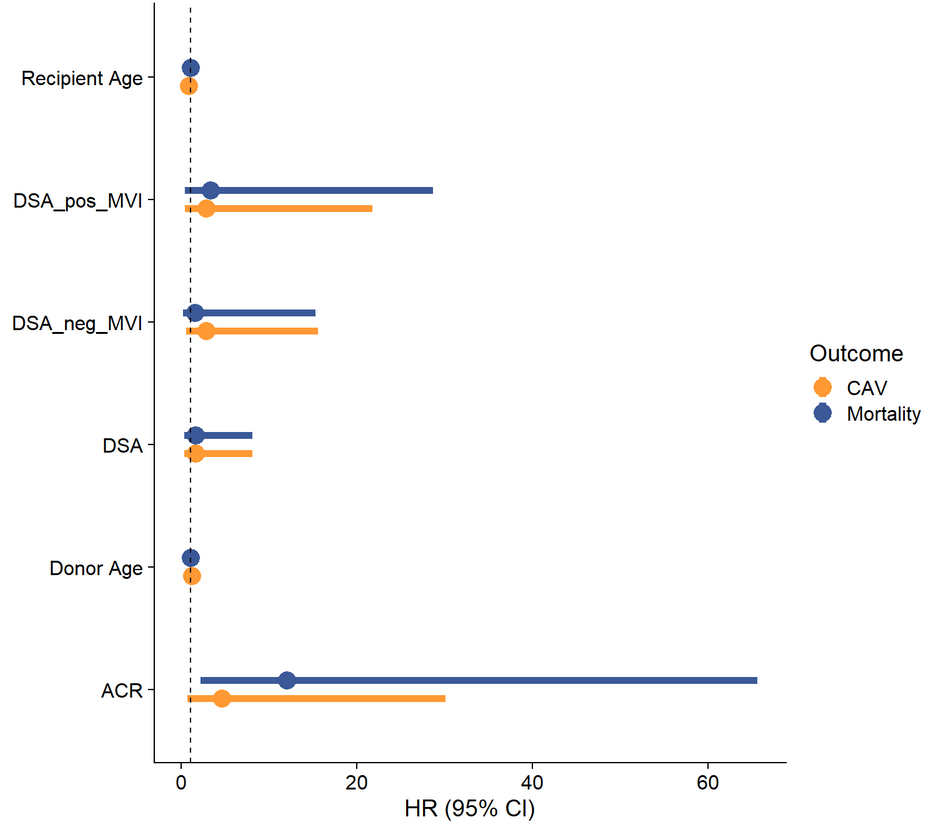

**Supplemental Figure 2.** Six-month landmark sensitivity analysis in full cohort (N = 832).

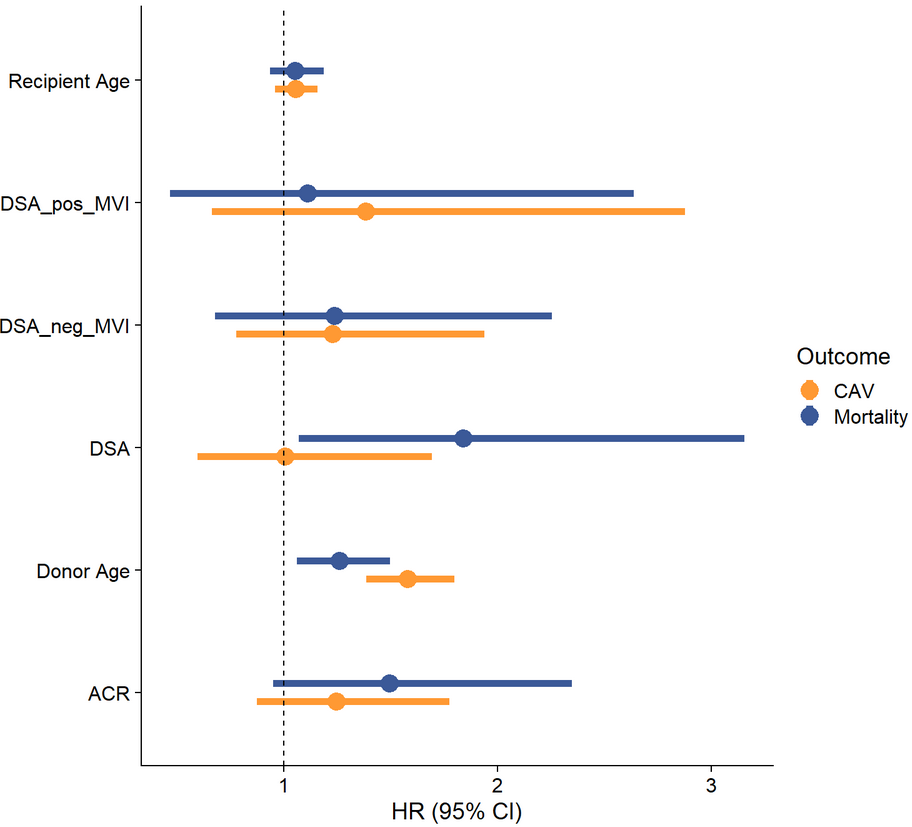
